# Laterality, Symmetry and Completeness Patterns of Nonsyndromic Orofacial Clefts in a Multiethnic Cohort

**DOI:** 10.64898/2025.12.29.25343144

**Authors:** Christina E. Spencer, Ligiane A Machado-Paula, Fang Qian, Azeez Butali, Carmen J. Buxo-Martinez, Carmencita D. Padilla, Claudia Restrepo-Muneton, Consuelo Valencia-Ramirez, Ross E. Long, Seth M. Weinberg, Mary L. Marazita, Jeffrey C. Murray, Lina M. Moreno Uribe, Aline L. Petrin

**Affiliations:** University of Iowa; University of Puerto Rico, School of Dental Medicine, Dental and Craniofacial Genomics Core; University of the Philippines; Clinica Noel, Medellín, Colombia; Lancaster, PA, Cleft Palate Clinic; University of North Carolina, Adams School of Dentistry; University of Pittsburgh, Center for Craniofacial and Dental Genetics, Department of Oral and Craniofacial Sciences, School of Dental Medicine; and Department of Human Genetics, School of Public Health

**Keywords:** Cleft Lip and Palate, Asymmetric Cleft, Laterality, Completeness, Unilateral Cleft Lip, Bilateral Cleft, Cleft Severity, LAHSHAL

## Abstract

**Objective:** Nonsyndromic cleft lip with or without cleft palate (NSCL/P) presents in diverse phenotypic forms defined by laterality, symmetry, and completeness. This study examined these features and their associations with sex, race, and ethnicity in a multiethnic cohort of 3,561 individuals from North America, South America, Asia, and Africa.

**Methods:** Associations were evaluated using Chi-square or Fisher’s exact tests with Bonferroni correction.

**Results:** Left unilateral clefts were the most prevalent, followed by right unilateral and bilateral clefts. The most common phenotype was a left complete unilateral cleft lip; the rarest was an asymmetric bilateral cleft with complete right-sided involvement. Among asymmetric bilateral cases, left-sided completeness was significantly more frequent than right-sided (p < 0.001), indicating the left side is both more commonly and more severely affected. Bilateral clefts — both symmetric and asymmetric — were more prevalent in males, while unilateral clefts were more common in females (p < 0.001). Asymmetric bilateral clefts occurred most frequently in Whites, whereas symmetric bilateral clefts were most common among Asians and Hispanics. Despite sex differences in bilateral cleft prevalence, completeness did not vary by sex for bilateral cases. For unilateral clefts, females showed higher completeness rates than males, and Asians and Hispanics/Latinos were more likely to exhibit complete unilateral clefts. Left unilateral clefts were most prevalent among Asians.

**Conclusions:** Sex, race, and ethnicity influence cleft laterality and symmetry, but cleft completeness appears sex-independent. The dissociation between side predominance and completeness suggests that distinct biological mechanisms govern cleft initiation and severity.

## Introduction

Nonsyndromic Orofacial Clefts (NS-OFC) affect approximately 1 in 700 individuals ^1,2^. The clinical presentation varies considerably, with cleft lip plus cleft palate representing approximately 45% of cases, isolated cleft palate 35%, and isolated cleft lip 20%. Unilateral cleft lip occurs more frequently than bilateral presentations, with a left-sided preponderance leading to a left:right:bilateral ratio of approximately 6:3:1^3–5^. Clefts of the lip with or without the palate affect males more often than females whilst cleft of the palate affect most commonly females ^6^ . Additionally cleft incidence also varies by ethnicity^5,7,8^. In this study we focus on Nonsyndromic Cleft Lip with or without Cleft Palate (NSCL/P).

Among bilateral cleft lip cases, both sides could present with complete clefts affecting the lip, alveolus, and nose (symmetric completeness) or, one side may present a complete cleft while the other side is incomplete (asymmetric completeness). The ratio of occurrence and etiology underlying symmetric versus asymmetric completeness in bilateral cases remains unknown. Similarly, in unilateral cases whether complete or incomplete presentations affect each side randomly, or preferentially occur on a particular side has not been established. After a thorough literature review, no consensus was found regarding the prevalence of right versus left completeness in asymmetric bilateral cases or on the distribution of completeness for unilateral cases; the available studies are summarized below.

Limited data exist on the prevalence of bilateral CL/P subtypes. Yuzuriha et al. reported that bilateral clefts present as symmetrical complete (54.4%), symmetrical incomplete (22.3%), and asymmetrical (23.3%) configurations^9^, while Rossell-Perry & Gavino-Gutierrez found that asymmetric cases accounted for 61.4% of their cohort^10^. Despite representing a substantial proportion of bilateral cases, asymmetric presentations —and particularly potential side preferences in completeness—remain largely uninvestigated.

While left-sided predominance in unilateral CL/P is well established, minimal research has been done into the completeness patterns for the unilateral clefts. Gatti et al. recorded the completeness of the unilateral clefts based on side. While they presented the number of patients, they did not look specifically at the prevalence and also included syndromic cases in their sample^11^. Thus, findings of a preferential side for complete clefting in unilateral cases also warrant further research.

Lastly, no studies have systematically compared completeness patterns between unilateral and bilateral presentations. This gap in knowledge limits our understanding of the developmental mechanisms underlying cleft formation and their relationship to embryological laterality establishment^12^.

Understanding laterality patterns in cleft completeness has important clinical implications. Bilateral cleft lip cases, particularly those with complete clefting, present greater surgical complexity and require more extensive reconstructive procedures to achieve optimal functional and aesthetic outcomes^13^. Accurate phenotypic characterization including a description of laterality and completion patterns can inform etiology, aid surgical planning and prognosis counseling^14^.

Laterality disorders can affect various organs and systems, including the heart, lungs, and abdominal organs. There are several factors that have been investigated to contribute to these disorders. During embryonic development, specific rotation of signaling molecules creates the gradient necessary for proper laterality establishment. Reversal of this rotation results in *situs inversus totalis*, where great vessels and thoracic and abdominal visceral organs develop on the opposite side from normal. While *situs inversus totalis* has complete mirroring, it is also possible to have incomplete mirroring. For instance, *situs abdominalis* has abdominal structures mirrored but not thoracic structures. Additionally, isomerism can be present in the heart or lungs. Isomerism of the heart presents with many other comorbidities, such as stenosis or communication of inappropriate compartments of the vascular system. ^14^ Although human laterality disorders—including craniofacial anomalies—have been investigated, the specific laterality patterns associated with complete and incomplete clefts or their specific etiology remain unclear.

This study aims to (1) determine the frequency of symmetric versus asymmetric completeness in bilateral clefts and identify any side predominance for completeness in asymmetric cases; (2) assess the frequency of complete versus incomplete presentations in unilateral clefts by side; (3) compare side-specific completeness patterns between bilateral and unilateral clefts to identify which side is overall most severely affected; and (4) examine associations between all cleft lip classifications including cleft completeness patterns and demographic variables, including sex, ethnicity, and race.

Our work addresses critical gaps in our knowledge of OFC presentations and provides groundwork for future investigations into the etiological mechanisms underlying complete versus incomplete defects and their relationship to laterality. Identification of preferential patterns of occurrence would warrant further research into the genetic and epigenetic factors contributing to these presentations.

## Materials and Methods

This retrospective cross-sectional study utilized previously collected data from four multiethnic cohorts across North and South America, Asia, and Africa. The study population comprised of 3,561 individuals with NSCL/P, including 76 cases with asymmetric bilateral clefts, 815 symmetric bilateral clefts and 2,670 cases with unilateral clefts. Cases lacking complete information on cleft type or completeness were excluded. To minimize familial confounding, only one family member was included per family unless the two individuals presented with different cleft subtypes (bilateral versus unilateral), as these represented distinct analytical subsets.

The LAHSHAL classification system was utilized when available to identify and characterize cleft presentations ^15^. For cohorts not utilizing the LAHSHAL system, data were analyzed and reclassified into a uniform classification system indicating cleft type and completeness to facilitate consistent sorting and statistical analysis. Bilateral cases were evaluated for asymmetry, defined as a complete cleft of the lip on one side and an incomplete cleft on the opposite side. Completeness was classified for both symmetric bilateral cases (i.e., symmetric complete or symmetric incomplete) and asymmetric bilateral cases, as well as for unilateral cases, to determine side predominance and characterize laterality patterns.

Demographic variables included sex, race, and ethnicity. Race originally was categorized into 4 groups (Asian, Black or African American, White, Other race). However, due to the small sample size of Asian patients and the apparent significance associated with race, it was deemed best to include a second race analysis where Asian patients were included in the Other race category. Data for specific cleft clinical characteristics were subdivided into several overlapping categories to provide greater analytic resolution, assist in the statistical analysis performed and facilitate quality checks for accurate case counts. These cleft clinical characteristics include cleft laterality, cleft completeness as well as cleft occurrence and severity by cleft side (Table 1).

**Table 1.** Patient Demographics and Clinical Cleft Characteristics by Cleft Lip Classification.

|  | **Total**<br>**N=3,561**<br>**n (%)** | **Asymmetric Bilateral**<br>**N=76**<br>**n (%)** | **Symmetric Bilateral**<br>**N=815**<br>**n (%)** | **Unilateral**<br>**N=2,670**<br>**n (%)** |
| --- | --- | --- | --- | --- |
| **Demographics:** |  |  |  |  |
| **Sex** |  |  |  |  |
| Female | 1,556 (43.7) | 27 (35.5) | 324 (39.7) | 1,205 (45.1) |
| Male | 2,004 (56.3) | 49 (64.5) | 491 (60.3) | 1,464 (54.8) |
| Unknown/Blank | 1 (0.0) | 0 (0.0) | 0 (0.0) | 1 (0.1) |
| **Race** |  |  |  |  |
| Asian | 450 (12.6) | 13 (17.1) | 125 (15.4) | 312 (11.7) |
| Black or African of American | 1,660 (46.6) | 24 (31.6) | 330 (40.5) | 1,306 (48.9) |
| White | 856 (24.1) | 29 (38.1) | 187 (22.9) | 640 (24.0) |
| Other Race | 591 (16.6) | 10 (13.2) | 172 (21.1) | 409 (15.3) |
| Unknown/Refuse to Answer | 4 (0.1) | 0 (0.0) | 1 (0.1) | 3 (0.1) |
| **Ethnicity** |  |  |  |  |
| Hispanic or Latino | 826 (23.2) | 12 (15.8) | 223 (27.4) | 591 (22.1) |
| Non-Hispanic or Latino | 2,404 (67.5) | 47 (61.8) | 515 (63.2) | 1,842 (69.0) |
| Unknown | 331 (9.3) | 17 (22.4) | 77 (9.4) | 237 (8.9) |
| **Clinical Cleft Characteristics:** |  |  |  |  |
| **Bilateral/Unilateral** |  |  |  |  |
| Bilateral | 891 (25.0) | 76 (100.0) | 815 (100.0) | 0 (0.0) |
| Right Unilateral | 1,000 (28.1) | 0 (0.0) | 0 (0.0) | 1,000 (37.5) |
| Left Unilateral | 1,670 (46.9) | 0 (0.0) | 0 (0.0) | 1,670 (62.5) |
| **Completeness of Lip Cleft** |  |  |  |  |
| Asymmetric, Left Complete | 56 (1.6) | 56 (73.7) | 0 (0.0) | 0 (0.0) |
| Asymmetric, Right Complete | 20 (0.6) | 20 (26.3) | 0 (0.0) | 0 (0.0) |
| Left Complete | 1,302 (36.6) | 0 (0.0) | 0 (0.0) | 1,302 (48.8) |
| Left Incomplete | 368 (10.3) | 0 (0.0) | 0 (0.0) | 368 (13.8) |
| Symmetric Complete | 734 (20.6) | 0 (0.0) | 734 (90.1) | 0 (0.0) |
| Symmetric Incomplete | 81 (2.3) | 0 (0.0) | 81 (9.9) | 0 (0.0) |
| Right Complete | 799 (22.4) | 0 (0.0) | 0 (0.0) | 799 (29.9) |
| Right Incomplete | 201 (5.6) | 0 (0.0) | 0 (0.0) | 201 (7.5) |
| **Right-Side Cleft** |  |  |  |  |
| Yes | 1,891 (53.1) | 76 (100.0) | 815 (100.0) | 1,000 (37.5) |
| No | 1,670 (46.9) | 0 (0.0) | 0 (0.0) | 1,670 (62.5) |
| **Right-Side Completeness** |  |  |  |  |
| Complete | 1,553 (43.6) | 20 (26.3) | 734 (90.1) | 799 (29.9) |
| Incomplete | 338 (9.5) | 56 (73.7) | 81 (9.9) | 201 (7.5) |
| NA/Left Unilateral Case | 1,670 (46.9) | 0 (0.0) | 0 (0.0) | 1,670 (62.6) |
| **Left-Side Cleft** |  |  |  |  |
| Yes | 2,561 (71.9) | 76 (100.0) | 815 (100.0) | 1,670 (62.5) |
| No | 1,000 (28.1) | 0 (0.0) | 0 (0.0) | 1,000 (37.5) |
| **Left-Side Completeness** |  |  |  |  |
| Complete | 2,092 (58.7) | 56 (73.7) | 734 (90.1) | 1,302 (48.8) |
| Incomplete | 469 (13.2) | 20 (26.3) | 81 (9.9) | 368 (13.8) |
| NA/Right Unilateral Case | 1,000 (28.1) | 0 (0.0) | 0 (0.0) | 1,000 (37.4) |
| **Side of Biggest Cleft** |  |  |  |  |
| Left | 1, 726 (48.5) | 56 (73.7) | 0 (0.0) | 1,670 (62.5) |
| Right | 1,020 (28.6) | 20 (26.3) | 0 (0.0) | 1,000 (37.5) |
| NA/Symmetric Case | 815 (22.9) | 0 (0.0) | 815 (100.0) | 0 (0.0) |

Data management was performed using Excel and statistical analyses were conducted using SAS® System version 9.4 (SAS Institute Inc., Cary, NC, USA). Descriptive statistics were used to summarize all variables of interest, including frequencies and corresponding percentages. Bivariate analyses were performed to examine associations between cleft lip classifications and demographic or clinical characteristics. Pearson’s chi-square test or Fisher’s exact test was applied as appropriate, with statistical significance set at α = 0.05. Chi-square goodness-of-fit test was used to evaluate laterality preponderances in bilateral asymmetric, and unilateral cases. Finally, cleft completeness as well as side preponderance and its associations with sex, ethnicity, and race were compared across phenotypic groups.

Because all analyses were hypothesis-driven and conducted within distinct clinical subgroups addressing different research questions, formal adjustments for multiple comparisons (e.g., Bonferroni correction or the Benjamini–Hochberg method) were not applied in the primary analyses. In observational epidemiologic studies with pre-specified hypotheses, strict multiple-comparison corrections are often not required, and their application may unnecessarily reduce statistical power and obscure clinically meaningful associations ^16–18^. However, because a total of 25 statistical tests were performed, a conservative Bonferroni correction (α = 0.05/25 = 0.002) was applied as a secondary sensitivity analysis to further assess the robustness of the observed associations.

## Results

A total of 3,561 patients with NS-CL/P were classified into three groups based on laterality and symmetry of cleft lip affection: asymmetric bilateral (n=76, 2.1%), symmetric bilateral (n=815, 22.9%), and unilateral (n=2,670, 75.0%). Table 1 presents the distribution of demographic and clinical cleft characteristics across these classifications for the total sample. Males comprised the majority of the sample (56.3%) compared to females (43.7%). Racial distribution in this multiethnic cohort showed that Black or African American patients constituted the largest group (46.6%), followed by White (24.1%), Other race (16.6%) and Asian (12.6%). In terms of Ethnicity, Non-Hispanics or Latino were the largest group (67.5%) followed by Hispanics (23.2%) and then by the Unknown ethnicity group (9.3%).

Regarding clinical cleft characteristics, left unilateral clefts (46.9%) were more common than right unilateral (28.1%) and bilateral clefts (25%). With respect to completeness, the most frequent presentation was a left complete cleft lip (36.6%), whereas the least common was an asymmetric bilateral cleft with a complete right side (0.6%). Overall, the left side of the lip is found affected more often and more severely than the right side.

### Patient Demographics and Cleft Lip Classification based on Laterality and Symmetry of Lip Affection

Table 2 shows results indicating that the risk of having an asymmetric bilateral, a symmetric bilateral or a unilateral cleft differed significantly by sex (p = 0.009), race (p < 0.001), and ethnicity (p < 0.001) for the overall cohort. Regarding sex, males showed higher risk for bilateral clefts overall, whereas females showed higher risk for unilateral clefts. Regarding race, the prevalence of asymmetric bilateral clefts ranged from 1.4% among Black or African American patients to 3.4% among White patients. Symmetric bilateral clefts were most common among individuals categorized as other race (29.1%) and Asians (27.8%), whereas unilateral clefts remained the most frequent classification across all racial groups (69.2%–78.7%). Similar patterns were observed when race categories were regrouped (p<0.001).

**Table 2.** Associations Between Patient Demographics and Cleft Lip Classification.

| Demographics | Cleft Lip Classification |  |  |  |
| --- | --- | --- | --- | --- |
|  | Asymmetric Bilateral<br>N=76<br>n (%) | Symmetric Bilateral<br>N=815<br>n (%) | Unilateral<br>N=2,670<br>n (%) | p-value |
| **Sex** |  |  |  | **0.009\*\*** |
| Female (n=1,556) | 27 (1.8) | 324 (20.8) | 1,205 (77.4) |  |
| Male (n=2,004) | 49 (2.5) | 491 (24.5) | 1,464 (73.0) |  |
| **Race** |  |  |  | **<0.001\*\*** |
| Asian (n=450) | 13 (2.9) | 125 (27.8) | 312 (69.3) |  |
| Black or African of American (n=1,660) | 24 (1.4) | 330 (19.9) | 1,306 (78.7) |  |
| White (n=856) | 29 (3.4) | 187 (21.8) | 640 (74.8) |  |
| Other Race (n=591) | 10 (1.7) | 172 (29.1) | 409 (69.2) |  |
| **Race (regrouped)** |  |  |  | **<0.001\*\*** |
| Black or African of American (n=1,660) | 24 (1.4) | 330 (19.9) | 1,306 (78.7) |  |
| White (n=856) | 29 (3.4) | 187 (21.8) | 640 (74.8) |  |
| Other Race (n=1,041) | 23 (2.2) | 297 (28.5) | 721 (69.3) |  |
| **Ethnicity** |  |  |  | **<0.001\*\*** |
| Hispanic or Latino (n=826) | 12 (1.5) | 223 (27.0) | 591 (71.5) |  |
| Non Hispanic or Latino (n=2,404) | 47 (2.0) | 515 (21.4) | 1,842 (76.6) |  |
| Unknown (n=331) | 17 (5.1) | 77 (23.3) | 237 (71.6) |  |

\*\*Statistically significant at $p < 0.05$ based on the chi-square test ( $p < 0.05$ ). Bonferroni correction $p < 0.002$

Lastly, regarding ethnicity, the proportion of asymmetric bilateral clefts ranged from 1.5% among Hispanic or Latino patients to 5.1% among those with unknown ethnicity. Further, symmetric bilateral clefts were most common amongst Hispanic or Latino (27%), while unilateral clefts remained the most common across all ethnic groups (71.5%–76.6%) with increase prevalence amongst Non Hispanics (76.6%).

### Cleft Completeness and Side Preponderance in Asymmetric Bilateral Cleft Lip

In the current study, 76 cases were classified as having an asymmetric bilateral cleft lip. Table 3 presents the distribution of patient characteristics associated with right-side completeness status. Patients were classified into two groups based on right-side cleft completeness: complete (n=20) and incomplete (n=56). Amongst the 76 patients with asymmetric bilateral cleft lip, left side completeness was significantly more frequent than the right side (73.7% vs. 26.3%; p<0.001). The chi-square goodness-of-fit test confirmed significant differences in the distribution of completeness for both right-side (30.5% complete vs. 69.5% incomplete; p=0.003) and left-side (69.5% complete vs. 30.5% incomplete; p=0.003) presentations indicating that in asymmetric bilateral cases a right-side complete affection is less prevalent; in other words the right side in asymmetric bilateral cases is often the less severely affected.

**Table 3.** Variables Associated with Right-Side Completeness Status among Patients With An Asymmetric Bilateral Cleft Lip (N=76)

| Variables | Right-Side Completeness Status |  |  |
| --- | --- | --- | --- |
|  | Complete (N=20)<br>n (%) | Incomplete (N=56)<br>n (%) | p-value* |
| **Left completeness** |  |  | **<0.001\*** |
| Complete (n=56) | 0 (0.0) | 56 (100.0) |  |
| Incomplete (n=20) | 20 (100.0) | 0 (0.0) |  |
| **Sex** |  |  | 0.302 |
| Female (n=27) | 9 (33.3) | 18 (66.7) |  |
| Male (n=49) | 11 (22.5) | 38 (77.5) |  |
| **Race** |  |  | 0.462 |
| Asian (n=13) | 4 (30.8) | 9 (69.2) |  |
| Black or African of American (n=24) | 7 (29.2) | 17 (70.8) |  |
| White (n=29) | 5 (17.2) | 24 (82.8) |  |
| Other Race (n=10) | 4 (40.0) | 6 (60.0) |  |
| **Race (regrouped)** |  |  | 0.336 |
| Black or African of American (n=24) | 7 (29.2) | 17 (70.8) |  |
| White (n=29) | 5 (17.2) | 24 (82.8) |  |
| Other Race (n=23) | 8 (34.8) | 15 (65.2) |  |
| **Ethnicity** |  |  | 0.088 |
| Hispanic or Latino (n=12) | 6 (50.0) | 6 (50.0) |  |
| Non Hispanic or Latino (n=47) | 12 (25.5) | 35 (74.5) |  |
| Unknown (n=17) | 2 (11.8) | 15 (88.2) |  |

\*Statistically significant using Fisher's exact test (p<0.05). Bonferroni correction p<0.002

Right-side completeness did not differ significantly by sex (p = 0.302), although females showed a numerically higher proportion of complete right-side cases compared with males (33.3% vs. 22.5%) this modest difference was not significant and indicates that sex does not predict right side-completeness in this sample.

Similarly, no significant associations were found between race and right-side completeness status (p=0.336), with racial distribution relatively balanced across completeness groups. Ethnicity showed a trend toward significance (p=0.088), with Hispanic or Latino patients more likely to have complete right-side clefts (50.0%) compared to non-Hispanic or Latino individuals (25.5%) and those with unknown ethnicity (11.8%).

### Cleft Completeness in Symmetric Bilateral Cleft Lip

Among the 815 patients with symmetric bilateral cleft lip, 734 (90.1%) presented with complete clefts and 81 (9.9%) with incomplete clefts. Chi-square goodness-of-fit testing showed a significant imbalance in completeness distribution, confirming a strong predominance of complete clefts in those affected with symmetric bilateral clefts (91.6% complete vs. 8.4% incomplete; p < 0.001).

Table 4 presents associations between demographic variables and completeness status. Sex was not significantly associated with completeness status (p=0.774), with similar rates observed among females (90.4%) and males (89.8%). Race was significantly associated with completeness (p<0.001), with White patients showing the lowest proportion of complete clefts (84.0%) compared to Asian (89.6%), Black or African American (90.3%), and Other Race (97.1%) patients. When race was analyzed in broader categories, the association remained significant (p=0.002), with White individuals consistently showing lower completeness rates.

**Table 4.** Variables Associated with Completeness Status among the Patients With A Symmetric Bilateral Cleft Lip (N=815)

| Variables | Right or Left Completeness Status |  |  |
| --- | --- | --- | --- |
|  | Complete (N=734)<br>n (%) | Incomplete (N=81)<br>n (%) | p-value |
| **Left completeness** |  |  | **<0.001\*** |
| Complete (n=734) | 734 (100.0) | 0 (0.0) |  |
| Incomplete (n=81) | 0 (0.0) | 81 (100.0) |  |
| **Sex** |  |  | 0.774 |
| Female (n=324) | 293 (90.4) | 31 (9.6) |  |
| Male (n=491) | 441 (89.8) | 50 (10.2) |  |
| **Race** |  |  | **<0.001\*\*** |
| Asian (n=125) | 112 (89.6) | 13 (10.4) |  |
| Black or African of American (n=330) | 298 (90.3) | 32 (9.7) |  |
| White (n=187) | 157 (84.0) | 30 (16.0) |  |
| Other Race (n=172) | 167 (97.1) | 5 (2.9) |  |
| **Race (regrouped)** |  |  | **0.002\*\*** |
| Black or African of American (n=330) | 298 (90.3) | 32 (9.7) |  |
| White (n=187) | 157 (84.0) | 30 (16.0) |  |
| Other Race (n=297) | 279 (93.9) | 18 (6.1) |  |
| **Ethnicity** |  |  | **<0.001\*\*** |
| Hispanic or Latino (n=223) | 215 (96.4) | 8 (3.6) |  |
| Non-Hispanic or Latino (n=515) | 461 (89.5) | 54 (10.5) |  |
| Unknown (n=77) | 58 (75.3) | 19 (24.7) |  |

\*Statistically significant using Fisher's exact test (p<0.05)
\*\* Statistically significant using chi-square test (p<0.05). Bonferroni correction p<0.002

Ethnicity was significantly associated with completeness status (p<0.001). Hispanic or Latino patients demonstrated the highest proportion of complete clefts (96.4%), followed by non-Hispanic patients (89.5%) and those with unknown ethnicity (75.3%).

### Cleft Completeness and Side Preponderance in Unilateral Cleft Lip

Among 2,670 patients with unilateral cleft lip, 2,101 (78.7%) had complete clefts and 569 (21.3%) had incomplete clefts. Chi-square goodness-of-fit testing confirmed a significant difference in completeness distribution (80.8% complete vs. 19.2% incomplete; p<0.001). Completeness status showed no significant association with cleft side (left vs. right; p=0.237), with complete presentations occurring in 78.0% of left-sided and 79.9% of right-sided clefts.

Table 5 presents associations between demographic variables and completeness status. Sex was significantly associated with completeness (p=0.003), with females showing higher rates of complete clefts (81.2%) compared to males (76.6%).

**Table 5.** Variables Associated with Cleft Completeness Status among the Patients with A Unilateral Cleft Lip (N=2,670)

| Variables | Cleft Completeness Status |  |  |
| --- | --- | --- | --- |
|  | Complete (N=2,101)<br>n (%) | Incomplete (N=569)<br>n (%) | p-value |
| **Side of Biggest Cleft**<br>Left (n=1,670)<br>Right (n=1,000) | 1,302 (78.0)<br>799 (79.9) | 368 (22.0)<br>201 (20.1) | 0.237 |
| **Sex**<br>Female (n=1,205)<br>Male (n=1,464) | 979 (81.2)<br>1,121 (76.6) | 226 (18.8)<br>343 (23.4) | **0.003\*\*** |
| **Race**<br>Asian (n=312)<br>Black or African of American (n=1,306)<br>White (n=640)<br>Other Race (n=409) | 265 (84.9)<br>1,061 (81.2)<br>440 (68.7)<br>334 (81.7) | 47 (15.1)<br>245 (18.8)<br>200 (31.3)<br>75 (18.3) | **<0.001\*\*** |
| **Race (regrouped)**<br>Black or African of American (n=1,306)<br>White (n=640)<br>Other Race (n=721) | 1,061 (81.2)<br>440 (68.7)<br>599 (83.1) | 245 (18.8)<br>200 (31.3)<br>122 (16.9) | **<0.001\*\*** |
| **Ethnicity**<br>Hispanic or Latino (n=591)<br>Non-Hispanic or Latino (n=1,842)<br>Unknown (n=237) | 497 (84.1)<br>1,469 (79.7)<br>135 (57.0) | 94 (15.9)<br>373 (20.3)<br>102 (43.0) | **<0.001\*\*** |

\*\* Statistically significant using chi-square test (p<0.05). Bonferroni correction p<0.002.

Race demonstrated a significant relationship with completeness (p<0.001). White patients had the lowest proportion of complete clefts (68.7%) compared to Asian (84.9%), Black or African American (81.2%), and Other Race (81.7%) individuals. This pattern remained significant when analyzed in broader racial categories (p<0.001).

Ethnicity was also significantly associated with completeness (p<0.001). Hispanic or Latino patients had the highest proportion of complete clefts (84.1%), while those with unknown ethnicity had the lowest (57.0%).

### Cleft side in Unilateral Cleft Lip Cases

Table 6 presents the association between demographic variables and cleft side among patients with unilateral cleft lip. Of 2,670 patients with unilateral cleft lip, 1,670 (62.5%) had a left-sided cleft and 1,000 (37.5%) had a right-sided cleft, confirming the established left-side predominance (p<0.001). Sex was marginally associated with cleft side (p=0.03) in unilateral clefts. Left-sided clefts occurred in 60.3% of females and 64.4% of males, while right-sided clefts occurred in 39.7% of females and 35.6% of males.

**Table 6.**
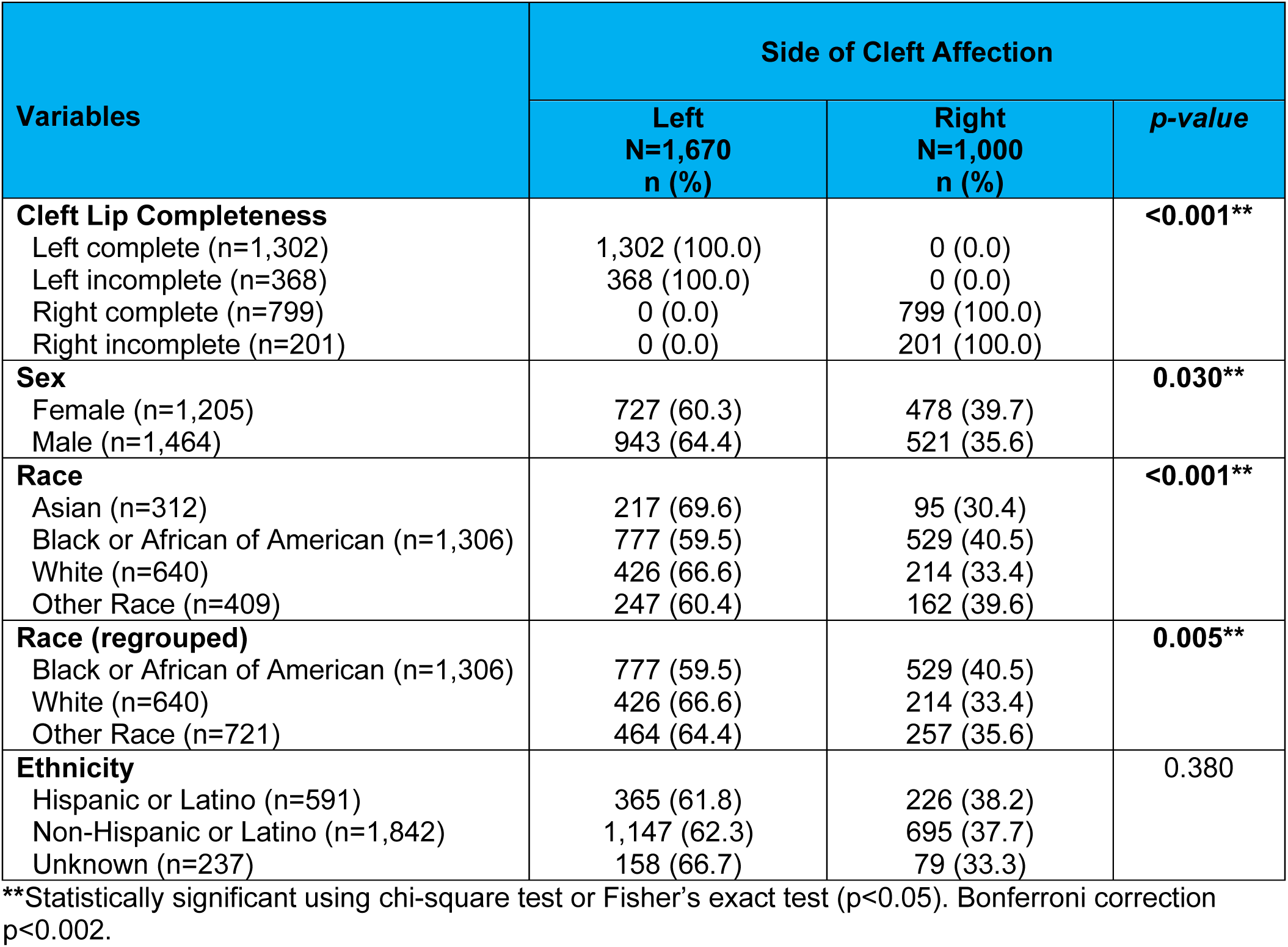
Association of Demographic and Clinical Variables with Cleft Side in Patients with Unilateral Cleft Lip (N = 2,670)

| Variables | Side of Cleft Affection |  |  |
| --- | --- | --- | --- |
|  | Left<br>N=1,670<br>n (%) | Right<br>N=1,000<br>n (%) | p-value |
| **Cleft Lip Completeness** |  |  | **<0.001\*\*** |
| Left complete (n=1,302) | 1,302 (100.0) | 0 (0.0) |  |
| Left incomplete (n=368) | 368 (100.0) | 0 (0.0) |  |
| Right complete (n=799) | 0 (0.0) | 799 (100.0) |  |
| Right incomplete (n=201) | 0 (0.0) | 201 (100.0) |  |
| **Sex** |  |  | **0.030\*\*** |
| Female (n=1,205) | 727 (60.3) | 478 (39.7) |  |
| Male (n=1,464) | 943 (64.4) | 521 (35.6) |  |
| **Race** |  |  | **<0.001\*\*** |
| Asian (n=312) | 217 (69.6) | 95 (30.4) |  |
| Black or African of American (n=1,306) | 777 (59.5) | 529 (40.5) |  |
| White (n=640) | 426 (66.6) | 214 (33.4) |  |
| Other Race (n=409) | 247 (60.4) | 162 (39.6) |  |
| **Race (regrouped)** |  |  | **0.005\*\*** |
| Black or African of American (n=1,306) | 777 (59.5) | 529 (40.5) |  |
| White (n=640) | 426 (66.6) | 214 (33.4) |  |
| Other Race (n=721) | 464 (64.4) | 257 (35.6) |  |
| **Ethnicity** |  |  | 0.380 |
| Hispanic or Latino (n=591) | 365 (61.8) | 226 (38.2) |  |
| Non-Hispanic or Latino (n=1,842) | 1,147 (62.3) | 695 (37.7) |  |
| Unknown (n=237) | 158 (66.7) | 79 (33.3) |  |

\*\*Statistically significant using chi-square test or Fisher's exact test ( $p < 0.05$ ). Bonferroni correction $p < 0.002$ .

Race was significantly associated with cleft side. In the original four-category classification, the highest proportion of left-sided clefts was observed among Asian patients (69.6%), followed by White patients (66.6%), Other race (60.4%), and Black or African American patients (59.5%) (p<0.001). When race was grouped into three categories (Black or African American, White, and Other race), the association remained statistically significant (p=0.005), with left-sided clefts occurring in 66.6% of White patients, 64.4% of patients in the Other race group and 59.5% of Black or African American patients.

Ethnicity showed no significant association with cleft side (p=0.380). Left-sided clefts occurred in 61.8% of Hispanic or Latino patients, 62.3% of non-Hispanic or Latino patients, and 66.7% of patients with unknown ethnicity.

## Discussion

This study investigated multiple classifications of lip involvement in NSCL/P—including laterality and symmetry of lip affection (i.e. Asymmetric bilateral, symmetric bilateral and unilateral clefts), cleft completeness in bilateral and unilateral clefts and side of cleft affection—in NSCL/P across a large, multiethnic cohort of 3,561 individuals. Overall, the distribution of cleft characteristics in this cohort aligns with established epidemiologic patterns of NSCL/P. Consistent with previous reports, left unilateral clefts were the most prevalent, followed by right unilateral clefts, with bilateral clefts being the least common. Regarding completeness, the most frequent presentation was a left complete unilateral cleft lip, whereas the rarest was an asymmetric bilateral cleft with complete right-side involvement. Collectively, these findings confirm that the left side of the lip is affected more frequently and more severely than the right.

We next examined associations of these lip classifications with demographic factors, including sex, race, and ethnicity. Across the full cohort, all three demographic factors are significantly associated with laterality and symmetry categories of lip affection (asymmetric bilateral, symmetric bilateral, and unilateral clefts) revealing distinct demographic patterns within these phenotypic categories.

In contrast, associations between demographic factors with cleft completeness were more variable. Notably, sex was not a risk factor for cleft completeness in bilateral clefts whether asymmetric or symmetric. Thus, although males are overall more frequently affected with bilateral clefts than females, both sexes appear to have a similar likelihood for developing complete versus incomplete bilateral clefts. Thus, the fact that complete cleft lip severity on bilateral clefts does not vary by sex indicates that the more extensive fusion failure is likely determined by timing (i.e. earlier occurrence for complete than incomplete clefts) and magnitude of neural crest–dependent developmental disruption, rather than sex-specific biological factors. In asymmetric bilateral clefts, race was not significantly associated with cleft completeness, while ethnicity showed only a marginal association with right-side completeness, with Hispanics exhibiting a slightly higher prevalence of complete right-side involvement. To our knowledge, this represents the first systematic evaluation of laterality in side-specific completeness among asymmetric bilateral clefts, including its association with demographic factors. The consistent predominance of left-side completeness regardless of sex, race, or ethnicity supports a stable left-sided directional pattern in cleft lip occurrence, even among asymmetric bilateral cases in which the right side appears comparatively more resistant to overt clefting. For symmetric bilateral clefts, both race and ethnicity emerged as significant risk factors, with Black individuals and Hispanics demonstrating the highest prevalence of complete symmetric bilateral clefts which are the most severe phenotypic affection.

Among individuals with unilateral clefts, cleft completeness did not exhibit a side preference; thus, although unilateral clefts occur more frequently on the left side than on the right overall, complete unilateral clefts occur at similar frequencies on either side^3,4,19^. Furthermore, unilateral cleft completeness was significantly associated with all three demographic factors.

However, consistent with findings in bilateral clefts above, sex showed only a marginal association with completeness risk, with females exhibiting a slightly higher likelihood of complete unilateral clefts than males indicating more severe phenotypic presentations in affected females. This aligns with previous studies^20^ that similarly observed greater completeness in females with unilateral NSCL/P. The higher severity in affected females may relate to sex-specific developmental factors. Interestingly, Hayashi et al. (1976) found that females with clefts matured earlier than males but exhibited greater underdevelopment in both maxillary and mandibular structures^21^. It is possible that the observed pattern of increased completeness in females reflect interactions between accelerated developmental timing and maxillary underdevelopment. As stated before, the male predominance for cleft occurrence observed in our overall cohort (56.3%) is consistent with established epidemiological patterns showing higher NSCL/P prevalence in males^7,8^. While females are less likely to be affected by cleft lip, our findings demonstrate that when affected, they tend to present with more severe phenotypes in unilateral cases. This sex-specific severity pattern warrants further investigation into the underlying biological mechanisms.

Regarding race and ethnicity complete unilateral clefts were significantly more common in Asians and Hispanics or Latino groups. Hispanic and certain racial minority groups appear predisposed to more severe cleft presentations when affected. These findings are particularly notable given that baseline cleft prevalence differs by race and ethnicity, with lower rates reported in Black and Hispanic populations.^22^ The observation that Hispanic and African populations present with more severe unilateral clefts despite lower overall prevalence suggests complex interactions between protective factors for cleft occurrence and factors influencing cleft severity. Interestingly, racial and ethnic disparities have been observed in other laterality-related conditions, such as breast cancer, where Hispanic and African American women show similar patterns in disease markers^23^, suggesting potential shared biological mechanisms worthy of exploration.

Finally, side of affection in unilateral clefts was strongly associated with race, with right-sided unilateral clefts more prevalent among Black individuals and left-sided unilateral clefts most common among Asians. Side of affection showed only a marginal association with sex and no association with ethnicity, suggesting that these factors are not strong predictors of left- versus right-sided occurrence in unilateral clefts.

Altogether, these findings provide important insights into the phenotypic diversity of NSCL/P and establish a foundation for future investigations into the developmental and genetic mechanisms underlying variation in cleft presentation.

### Biological implications

The study of cleft laterality and symmetry, completeness and preponderance on side of affection has implications for discovering etiological mechanisms such as variable expressivity, penetrance differences, modifiers genes and epigenetic risk factors that shape our understanding of the full phenotypic spectrum of orofacial clefting anomalies. In regard to the etiological mechanisms underlying asymmetric cleft patterns, while other human laterality disorders, such as *situs inversus totalis* are associated to the reversal of signaling molecule gradients resulting in complete body laterality reversal^14^, for NSCL/P, it is plausible that localized asymmetric disruptions in developmental signaling pathways contribute to the observed asymmetric cleft patterns. The interplay of genetic and environmental factors in laterality disorders, as demonstrated in heterotaxia^24^, suggests that multiple factors also contribute to the laterality patterns observed in asymmetric bilateral clefts. Like other laterality disorders, genetic and environmental factors both play significant parts in leading to a malformation and isolating each factor’s independent contribution is challenging.^25^ This represents a promising area for future research into the development of craniofacial laterality disorders.

Regarding preferential side for cleft affection, there have been studies showing overall systemic differences in cases that are correlated with the side of the cleft. For instance, Al-Hassani et. al found that individuals with right sided clefts had poorer speech and hearing outcomes while those with left clefts were more likely to be teased. They hypothesize that the poorer outcomes of the right cleft affected individuals have to do with the previously mentioned finding that patients with right unilateral CL/P have a greater degree of lateral lip element hypoplasia leading to greater deficiencies of lateral lip element vertical and vermillion. ^26^ Established associations have also been reported between unilateral CL/P sidedness and handedness, as well as congenital dental abnormalities. In addition, several studies have shown that right-sided unilateral CL/P is associated with a higher prevalence of congenital anomalies, underscoring that cleft sidedness is not an isolated phenotypic feature. Thus, cleft sidedness appears to be linked with distinct comorbidities, as evidenced by the correlations identified to date. These findings suggest important implications for etiologic research and indicate that, at a minimum, right- and left-sided unilateral clefts should be analyzed as separate entities.

### Clinical Implications

As knowledge of the biological mechanisms underlying laterality and cleft completeness continues to advance, our may have important clinical implications—particularly if differences in surgical outcomes between unilateral right-sided and left-sided clefts are confirmed. Such differences may be related to the observation that the right side clefts are often characterize by greater lip deficiencies likely impacting treatment approaches and outcomes ^26^. Moreover bilateral CL/P (NS-OFC) cases, particularly those with complete presentations, require more extensive reconstructive procedures to achieve optimal functional and aesthetic outcomes^12^. As emphasized by Soofi et al., accurate phenotypic characterization of laterality disorders is essential for optimal diagnosis and treatment^14^. Recognizing that certain demographic and ethnic groups may be more likely to present with complete clefts can also help clinicians anticipate surgical complexity and provide families with more tailored counseling regarding expected interventions and outcomes. Finally, the demographic associations identified in this study may inform risk stratification and support effective resource allocation for complex cleft care.

### Limitations

Some limitations should be considered when interpreting these findings. First, the retrospective design and reliance on existing databases may introduce selection bias, particularly given the overrepresentation of certain racial groups relative to population-based cleft prevalence. Second, the relatively small number of asymmetric bilateral cases (n=76) limited statistical power for detecting associations within this subgroup, particularly for the ethnicity analysis that approached but did not reach significance. Finally, this study focused on cleft lip completeness and did not systematically analyze palate involvement patterns, which warrants future investigation as well.

### Future Directions

This research establishes important groundwork for future mechanistic studies. The identification of laterality preponderances in asymmetric bilateral cases and demographic associations with cleft severity points toward potential genetic, epigenetic, and environmental factors worthy of investigation. Future research should explore the developmental biology underlying laterality establishment in craniofacial structures, examine gene-environment interactions that may contribute to observed racial and ethnic differences, and investigate the mechanisms underlying sex-specific severity patterns. Additionally, expanded cohorts with adequate representation of Hispanic and other minority populations are needed to definitively establish ethnic-specific patterns. Integration of genetic data with phenotypic characterization will be essential for elucidating the etiological mechanisms underlying the diverse presentations of NS-OFC. Future studies could also determine if there are outcome differences based on sex or race specific patterns that might alter surgical approaches or timing.

Understanding the factors that determine whether a cleft occurs as well as its laterality and severity represents a key step toward personalized medicine approaches utilized in cleft care. The molecular pathways controlling laterality in other organ systems, such as those involving signaling molecule gradients^14^ and the genetic contributors to heterotaxia^25,27^, provide promising avenues for investigation in the craniofacial context.

## Conclusions

This large multiethnic study provides the first systematic documentation of laterality preponderances in asymmetric bilateral NS-OFC, demonstrating significant left-side predominance for completeness. While unilateral clefts show well-established left-sided occurrence predominance, completeness appears independent of side in these cases. Importantly, we identified significant associations between cleft completeness and demographic factors, with females, non-White racial groups, and Hispanic ethnicity associated with more severe (complete) presentations. These findings enhance our understanding of NS-OFC phenotypic diversity and provide a foundation for future investigations into the genetic, epigenetic, and environmental mechanisms underlying cleft laterality and severity. The demographic associations identified may also inform clinical practice through improved risk stratification and surgical planning.

## Acknowledgments

We extend our sincere gratitude to the families who generously contributed clinical data to this study. We also thank the University of Iowa Student Research Program for providing the opportunity and additional financial support for this research.

## Ethical approval statement

The study was approved by the University of Iowa institutional review board (IRB# 202305391). All data used were obtained from previous IRB approved studies.

## Conflict of Interest

The authors declared no potential conflicts of interest with respect to the research, authorship, and/or publication of this article.

## Funding

This work was supported by the National Institutes of Health NIDCR NIH [U01DE024425, R21DE016930, R37 DE008559, R01DE011931, R00DE024571, R01DE016148, R01 DE014667], NCBDD CDC [R01DD000295], NIMHD NIH [S21MD001830], and the University of Iowa Department of Orthodontics, and College of Dentistry Student Research Program.

## Data availability statement

All data generated in this study is presented in the manuscript.

## Notes

### Competing Interest Statement

The authors have declared no competing interest.

### Summary of Updates

Revised text and statistical analysis.

